# Compounded environmental health risks in mountain communities upstream of Bhumibol Dam, Thailand

**DOI:** 10.64898/2026.01.09.26343813

**Authors:** Poramad Trivalairat, Isara Phiwchai, Matika Chaichan, Narisara Sripo

**Affiliations:** Princess Agrarajakumari Faculty of Nursing, Chulabhorn Royal Academy, 906 Thung Song Hong, Lak Si, Bangkok, 10210, Thailand; Laboratory for Exploring the Ectoparasites and Carnivorous Hirudineans (L.E.E.C.H.), Department of Zoology, Faculty of Science, Kasetsart University, 50 Ngam Wong Wan Road, Chatuchak, Bangkok, 10900, Thailand

**Author notes:** These authors contributed equally to this work.

**Keywords:** sand-filtered puddle, water quality, drinking water, contamination, mountain area, pesticide, urolithiasis

## Abstract

Indigenous, mountain communities residing upstream of Bhumibol Dam, Thailand, rely on vulnerable natural water sources for their water supply, yet remain unaware of the associated health risks. This study assessed the water quality, usage patterns and contamination pathways across six villages upstream of Bhumibol Dam to shed light on the obstacles to sustainable water security . Samples from 38 water sources of drinking and/or non-drinking water, soil, and the edible parts of crops were subjected to analyses of physical, chemical (NO_3_-N, pH), and qualitative pesticide-related variables, alongside a 6-month assessment of a community water filter system. Principal component analysis identified a “at-risk group” of preferred drinking water sources all exhibiting high NO_3_-N, highly alkaline pH, and substantial pesticide contamination, which was found to likely be caused by agricultural run-off. This was reinforced by the detection of pesticide residues in all soil samples and, critically, in the below-ground edible parts of crops (taro, lemongrass, arrowroot), confirming dietary exposure in the local communities. Further compounding the risks posed by the unsafe water supply, the community water filter was found to be ineffective throughout the 6-month analysis with there being no significant difference in water quality between before and after filtration. The residents’ paradoxical preference for high-risk, still water (from sand-filtered puddles) for drinking, rather than water from flowing sources, which they used only for cooking and cleaning. These findings reveal a severe, compounded public health threat of chronic exposure to minerals linked to urolithiasis and agrochemicals, highlighting the urgent need for quantitative risk assessment and the implementation of resilient, decentralized water treatment solutions in these mountain communities.

## Introduction

Access to clean water is essential for human health, supporting hydration and overall body function, as well as enabling hygienic cooking and cleaning practices. According to the principles of the Ottawa Charter and the tenets upheld by the World Health Organization (WHO), access to clean drinking water is a fundamental component of public health and a basic human right that promotes health equity and well-being [1,2]. In rural areas, particularly those at high altitudes, communities depend on water sources derived from mountain for their daily life. This water is renowned for its purity and high mineral content, which can help to maintain balanced pH and support hydration, while also being essential for agriculture. However, the high levels of alkaline minerals such as calcium and magnesium in water in mountainous regions can also lead to health complications, including the formation of bladder stones [3–7]. Moreover, many rural communities around the world are facing challenges to their water security exacerbated by contamination (with herbicides, pesticides, and heavy metals) and climate change (such as long droughts and less precipitation), which has influenced the availability of clean fresh water. This necessitates the development of effective water management strategies to ensure that populations continue to have safe access to this vital resource [3,8–11]. Indeed, the responsible stewardship of mountain water is critical for safeguarding the well-being of mountain communities and securing their sustainable access to clean drinking water.

Bhumibol Dam, the second-largest dam in Thailand, was constructed in 1953 and is located within the Mae Tuen Wildlife Sanctuary in Sam Ngao District, Tak Province, northwest Thailand. Currently, almost 2,000 members of the Paganyaw (S’gaw Karen) ethnic group live in mountain communities located above the dam. Given the long distances to the dam and its reservoir and the difficulty of traveling through this mountainous region, these communities’ water supply relies in part on systems derived water from mountain stream developed through community collaboration as part of a rural development program. While preliminary testing of the quality of drinking water from these systems was shown to be superior to that of natural sources, subsequent, more thorough testing revealed contamination. Specifically, water samples were found to breach safe drinking standards via the presence of 260 MPN/100 mL (MPN: most probable number) of coliform bacteria (including 7 MPN/100 mL of fecal coliform bacteria). Moreover, some residents of this region are unable to access the improved drinking water provided by these systems due to their remoteness, forcing them to consume untreated water from natural sources, particularly from sand-filtered puddles near streams [12–17]. This exposes them to various waterborne diseases, including acute abdominal pain, diarrhea, gastrointestinal conditions, and urinary issues, linked to contaminated water [18–20]. Moreover, widespread agricultural practices in this area, especially monoculture farming, exacerbate these health risks because of the intensive use of pesticides, which contaminate local water supplies and enter the food chain via fish. Given the remote geographical location, lack of reliable telecommunications and electricity, and limited access to healthcare in these vulnerable communities, there is an urgent need to improve water quality and sanitation.

The above background highlights the need to comprehensively assess the water sources used by mountain communities upstream of Bhumibol Dam in terms of their water quality and contamination levels. Such assessment is essential for effective management of the water supply for rural residents who live far from modern water infrastructure. Accordingly, this study was implemented with the aims of characterizing the quality of natural drinking water sources and water storage facilities used by the mountain communities upstream of Bhumibol Dam; investigating the pathways of agrochemical contamination into water, soil, and crops; and assessing the effectiveness of community water filtration systems. The overarching goal of this work is to enhance sustainable access to safe drinking water and improve health outcomes for these vulnerable communities.

## Materials and methods

### Study location and water sources

Mountain communities upstream Bhumibol Dam have been divided into six discrete “villages” since the building of the dam within Mae Tuen Wildlife Sanctuary, Ban Nar Sub-district, Sam Ngao District, Tak Province, northwest Thailand (Fig. 1): Aum Haum (V1), Aum Vab (V2), Hin Lat (V3), Nahi (V4), Nam Sub (V5), and San Pa Puai (V6). The location of each village was recorded and plotted on a map using QGIS (v.3.44), with subsequent editing of legends and additional information using Adobe Photoshop (v.13.0). Given the absence of tap water, residents of these villages rely on water sources such as waterfalls, rivers, canals, rainwater, or sand-filtered puddles. They use the water from these sources directly or store it in containers in their homes. In this study, water samples were collected from various water resources used by local residents across these six villages between 19 and 21 November, 2024, to assess water quality. Thirty-eight water sources were purposively selected as they represented the total land area accessible to and used by the community at the time of data collection. This approach was chosen to ensure comprehensive coverage, given the geographical dispersion and limited number of water sources available in these remote villages.

**Fig. 1.**
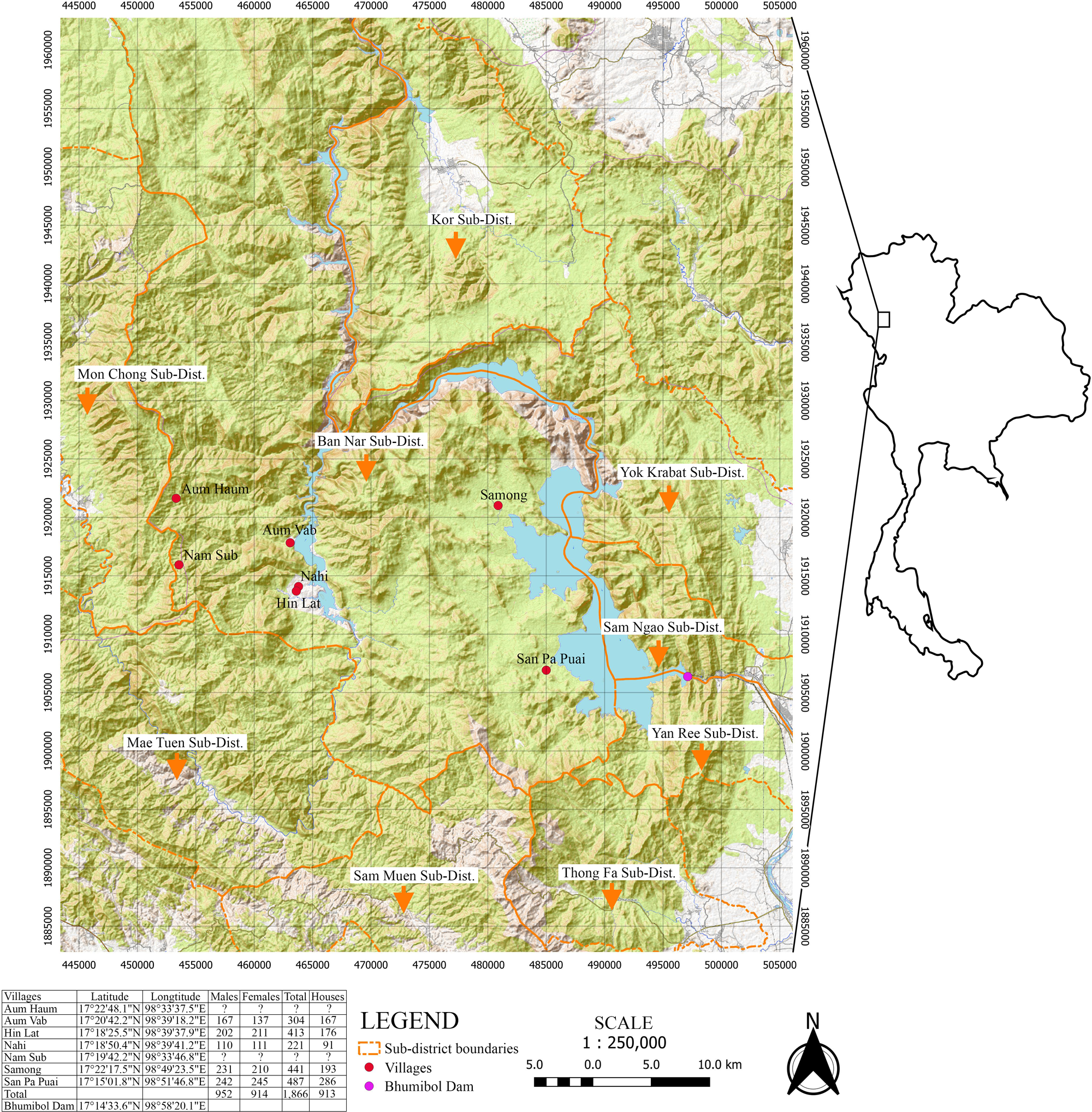
Map of villages upstream of Bhumibol Dam, Sam Ngao District, Tak Province, Thailand.

Water status at each site was categorized based on its use and characteristics, including: (1) source (SOURCE)—natural source vs. stored water (in containers); (2) drinking status (DRINK)—water used for drinking (drinking water) vs. water used only for other daily activities (non-drinking water) ; and (3) flow of the water (FLOW)—flowing vs. still. These factors were recorded using the YSI EXO multiparameter instrument (YSI Inc., Yellow Springs, OH, USA), along with physical parameters including height above sea level (ASL, m), specific conductivity (SPC, μs/cm), nitrate nitrogen (NO_3_-N) (NO3N, mg/l), mass of dissolved oxygen (DOm, mg/l), percentage of dissolved oxygen (DOp, %), pH, salinity (SAL, ppt), total dissolved solids (TDS, mg/L), and temperature (TEMP, °C). The data were subsequently analyzed to explore the relationships between water sources, environmental conditions, and water uses among the residents of each village.

Water samples were also collected from a single community water filter at Hin Lat village, which filters water from a mountain stream and supplies it to each household. The maintenance records of this community filtration system indicated that washing and filter replacement were infrequently performed, suggesting that its performance was suboptimal. Samples were collected from both the pre-filtration and post-filtration tanks of this system, in line with the sampling procedures outlined in the Standard Methods for the Examination of Water and Wastewater [21] for assessing the quality of drinking water. The samples were then separately collected into three sterile polyethylene bottles for laboratory analysis of physical and chemical indicators of water quality, microbial contamination, and heavy metals (Table 1). An acid-washed sterile bottle was used specifically for heavy metal analysis to prevent contamination and preserve metal ions. Samples were stored at 1–4°C and transported immediately to the Public Health Laboratory Division, Department of Health, Ministry of Public Health, Thailand, within 24 h for further examination. Furthermore, water samples from this filter were tested and assessed three times over a 6-month period with 3-month intervals to ensure consistency and reliability of the results.

**Table 1.**
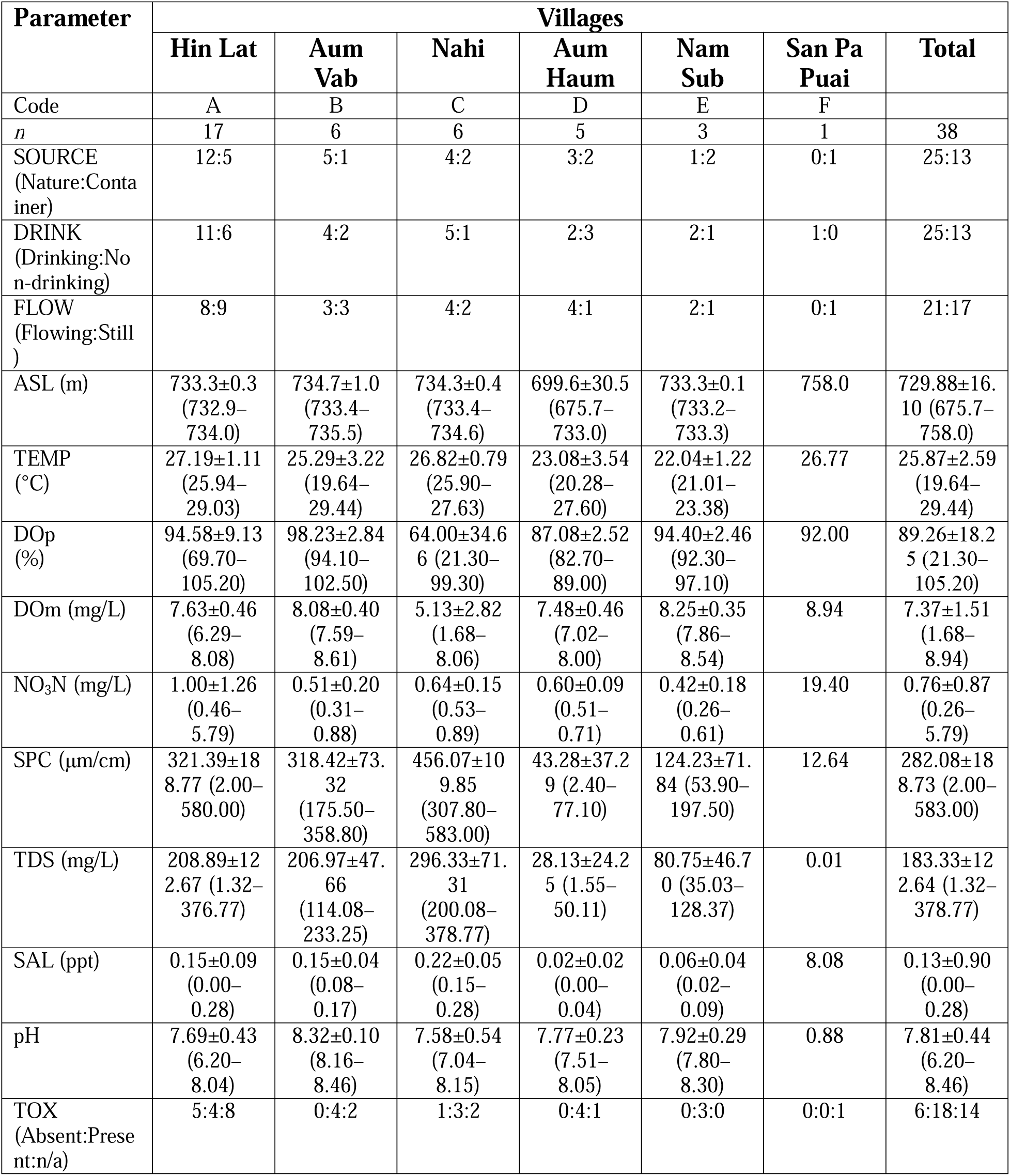
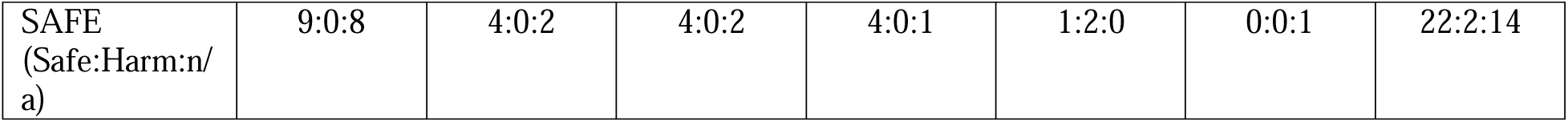
Water quality parameters analyzed across villages upstream of Bhumibol Dam, Sam Ngao District, Tak Province, Thailand, on 19 November, 2024. Abbreviations: ASL = height above sea level; TEMP = temperature; DOp = percentage of dissolved oxygen; DOm = mass of dissolved oxygen; NO3N = nitrate nitrogen; SPC = specific conductivity; TDS = total dissolved solids; SAL = salinity; TOX = presence of toxic substances (herbicides, insecticides); SAFE = water safety level for utilization.

### Pesticide analysis

To assess potential pesticide exposure and associated risk factors, samples of the edible part of crops, soil, and drinking water were collected and tested for the presence and level of pesticides. Specifically, the testing of targeted pesticides from the organophosphate, carbamate, and cholinesterase inhibitor groups was performed using the GT-Pesticide Residual Test Kit (Bsmartscience Co., Ltd., Mueang Nonthaburi, Nonthaburi, Thailand). This kit extracts pesticides from water, soil, and agricultural products and indicates their presence or absence (TOX) by comparing the colour of the extracted sample tube with that of a control tube. A colour that is lighter than or similar to that of the control indicates the absence of pesticides (score 0), while a darker colour suggests their presence (score −1). Additionally, the kit provides qualitative assessment of the toxicity levels (SAFE). Specifically, by comparing the sample’s colour with that of a standardized comparison tube, a lighter colour indicates that the pesticide is present but within a safe level (score 0), whereas a darker colour indicates the presence of pesticides at an unsafe level (score 1).

### Statistical analysis

Normality tests conducted using the Shapiro–Wilk method indicated that most water quality parameters did not follow a normal distribution within each village (p < 0.05). Therefore, non-parametric statistical methods were employed, with Kruskal–Wallis tests conducted in SPSS 27.0 to compare differences among villages. To identify specific pairwise differences, Mann–Whitney U tests were performed as post hoc analyses. These tests were also applied to data collected from the water filter at Hin Lat at three different timepoints to assess the consistency and reliability of the water quality measurements over time.

In addition to the non-parametric tests, Pearson’s or Spearman’s correlation analyses were conducted to examine the relationships between water status (SOURCE, DRINK, FLOW) and water parameters (SPC, NO_3_N, DOm, DOp, pH, SAL, TDS, TEMP, TOX, SAFE). The choice between Pearson’s and Spearman’s correlation depended on the data distribution. Spearman’s rank correlation was used for non-normally distributed variables.

### Multivariate analysis

Principal component analysis (PCA) was performed using Python libraries and dependence was analysed via the Google Colab environment, allowing exploration and description of the relationships among various water status and water quality parameters. Variables included water status (SOURCE, DRINK, FLOW), physical parameters (SPC, NO3N, DOm, DOp, pH, SAL, TDS, TEMP), and toxicity indicators (TOX, SAFE). PCA helped to identify underlying patterns and key factors influencing water quality and usage across the study sites.

### Ethics statement

This research was approved by the Human Research Ethics Committee, Chulabhorn Royal Academy (approval number: EC 030/2567).

## Results

### Water use

This study assessed water quality across six villages located upstream of Bhumibol Dam in Sam Ngao District, Tak Province, between 19 and 21 November, 2024. The villages spanned an altitudinal range of 675.7 to 758.0 m ASL (mean 729.88±16.10). A total of 38 water sources utilized by residents were sampled and analyzed for water status and various physical parameters as follows: Aum Haum (V1, n = 5), Aum Vab (V2, n = 6), Hin Lat (V3, n = 17), Nahi (V4, n = 6), Nam Sub (V5, n = 3), and San Pa Puai (V6, n = 1). The results indicated that 13 sources of natural flowing water were primarily used for non-drinking purposes (such as cleaning and cooking) (Fig. 2). The other 25 sources were used for drinking, with residents most commonly showing a preference for obtaining their drinking water from natural still water (sand-filtered puddles) from 10 sources, still water stored in containers (jars or concrete tanks) from 7 sources, flowing water derived through pipeline from mountain stream from 6 sources, and natural flowing water from 2 sources (Fig. 3). Regarding water status, the residents generally preferred natural water over water in containers (25:13) and flowing water over still water (21:17) for drinking purposes (25:13). However, the results of independent sample *t*-tests indicated that there was no difference in water use between each village for the parameters SOURCE, DRINK, and FLOW (*p*-values 0.562, 0.935, and 0.907, respectively).

**Fig. 2.**
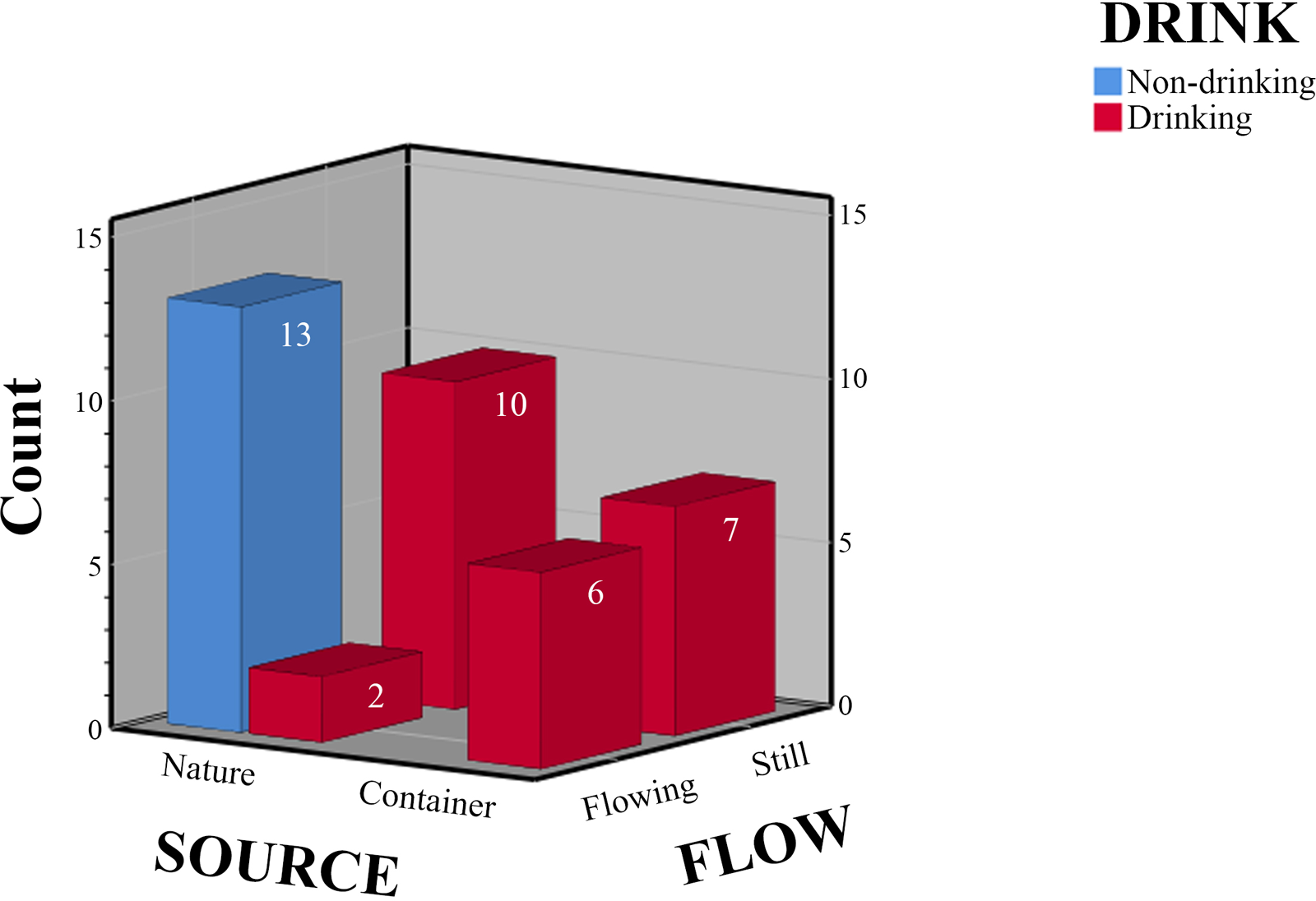
A 3D cluster bar chart displaying the uses of water sources, categorised by sources (SOURCE), flow condition (FLOW), and drinking status (DRINK) across six villages located upstream of Bhumibol Dam, between 19 and 21 November, 2024.

**Fig. 3.**
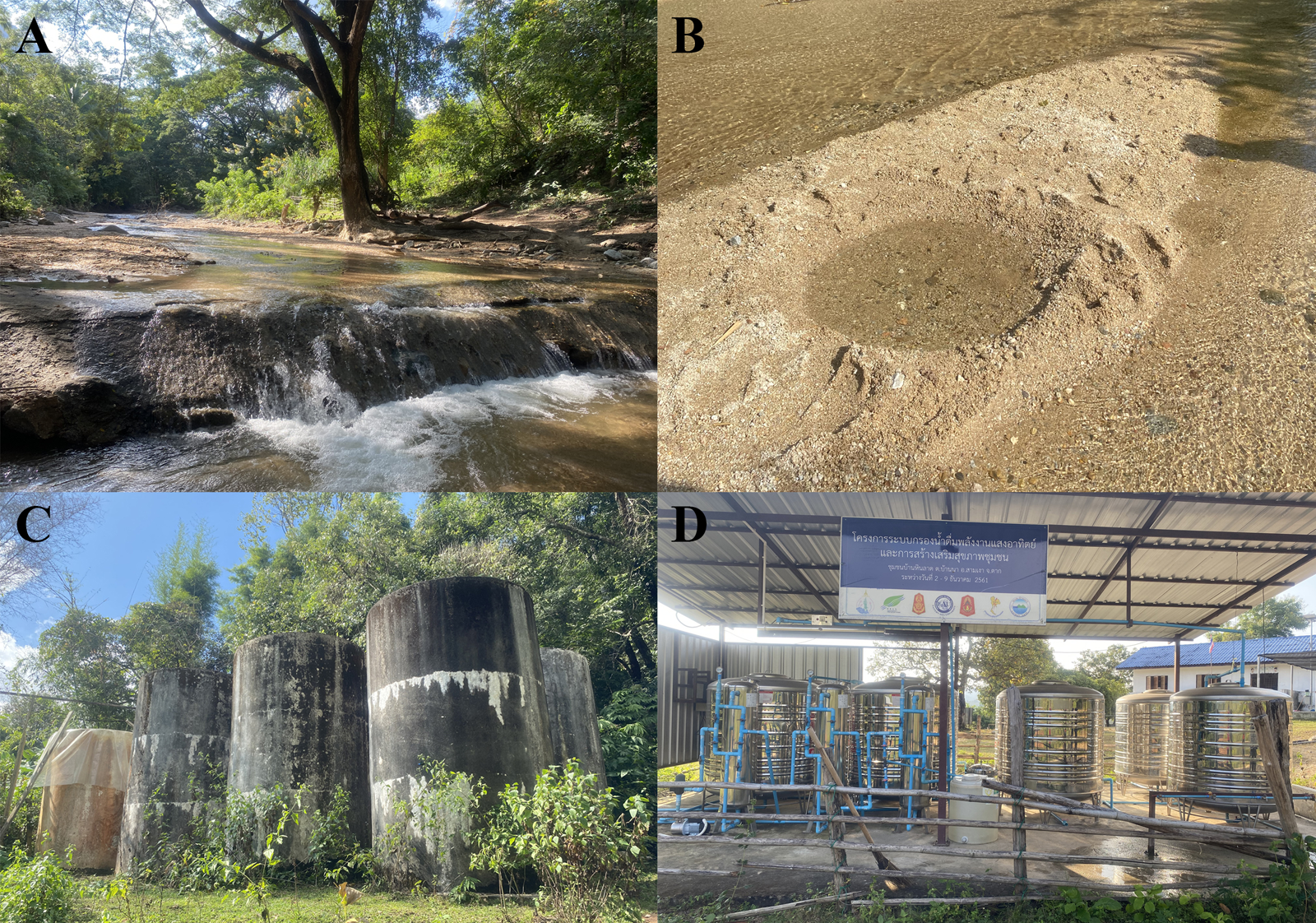
Sources of water used by residents upstream of Bhumibol Dam, including (A) mountain river in Aum Vab; (B) sand-filtered puddle in Aum Vab; (C) concrete tanks for storing water drained from waterfalls in Aum Haum; and (D) water filter tanks in Hin Lat.

### Water quality

Across the six villages, the water quality parameters were as follows: mean TEMP, 25.87±2.59°C (range 16.77–29.44°C); mean DOp, 89.26±18.25% (range 21.30%–105.20%); mean DOm, 7.37±1.51 mg/L (range 1.68–8.94 mg/L); mean NO_3_N, 0.76±0.87 mg/L (range 0.26–5.79 mg/L); mean SPC, 282.08±188.73 μs/cm (range 2.00–583.00 μs/cm); mean TDS, 183.33±122.64 mg/L (range 1.32–378.77 mg/L); mean SAL, 0.13±0.90 ppt (range 0.00–0.28 ppt); and mean pH, 7.81±0.44 (range 6.20–8.46) (Table 1).

Normality tests (Shapiro–Wilk) revealed that most water quality parameters were not normally distributed within each village (p < 0.05). Therefore, non-parametric statistical methods were used. Kruskal–Wallis tests revealed significant variations in various water quality parameters across the six villages surveyed. Specifically, statistically significant differences were observed for TEMP (χ^2^ = 12.558, p = 0.028), DOp (χ^2^ = 13.805, p = 0.017), DOm (χ^2^ = 15.189, p = 0.01), χ SPC (χ^2^ = 16.905, p = 0.005), TDS (χ^2^ = 16.903, p = 0.005), SAL (χ^2^ = 17.079, p = 0.004), and pH (χ^2^ = 16.343, p = 0.006) (Table 1). No significant differences were detected for NO N χ across the villages.

To identify specific differences between the villages, post hoc Mann–Whitney U tests were performed. These analyses indicated significant differences in TEMP between V1 and V3 (p = 0.031), V3 and V6 (p = 0.007), and V4 and V6 (p = 0.020). For DOp, significant differences were found between V1 and V2 (p = 0.006), V1 and V3 (p = 0.025), V1 and V6 (p = 0.024), V2 and V4 (p = 0.025), and V3 and V4 (p = 0.027). Significant differences in DOm were observed between V2 and V3 (p = 0.042), V2 and V4 (p = 0.025), V3 and V6 (p = 0.021), and V4 and V6 (p = 0.030). Regarding SPC, significant differences occurred between V1 and V2 (p = 0.006), V1 and V3 (p = 0.014), V1 and V4 (p = 0.006), V2 and V6 (p = 0.039), and V4 and V6 (p = 0.020). “Similarly, TDS, SAL, and SPC showed significant differences between villages (see Table 2). Significant differences in pH were found between V1 and V2 (p = 0.006), V2 and V3 (p < 0.001), V2 and V4 (p = 0.004), and V2 and V6 (p = 0.038). Moreover, NO_3_N and SAFE did not show significant differences across villages, except for between V2 and V3 for NO3N (p = 0.039) and between V3 and V6 for SAFE (p = 0.010). No other statistically significant differences were observed between the remaining pairs of villages for the parameters examined.

**Table 2.**
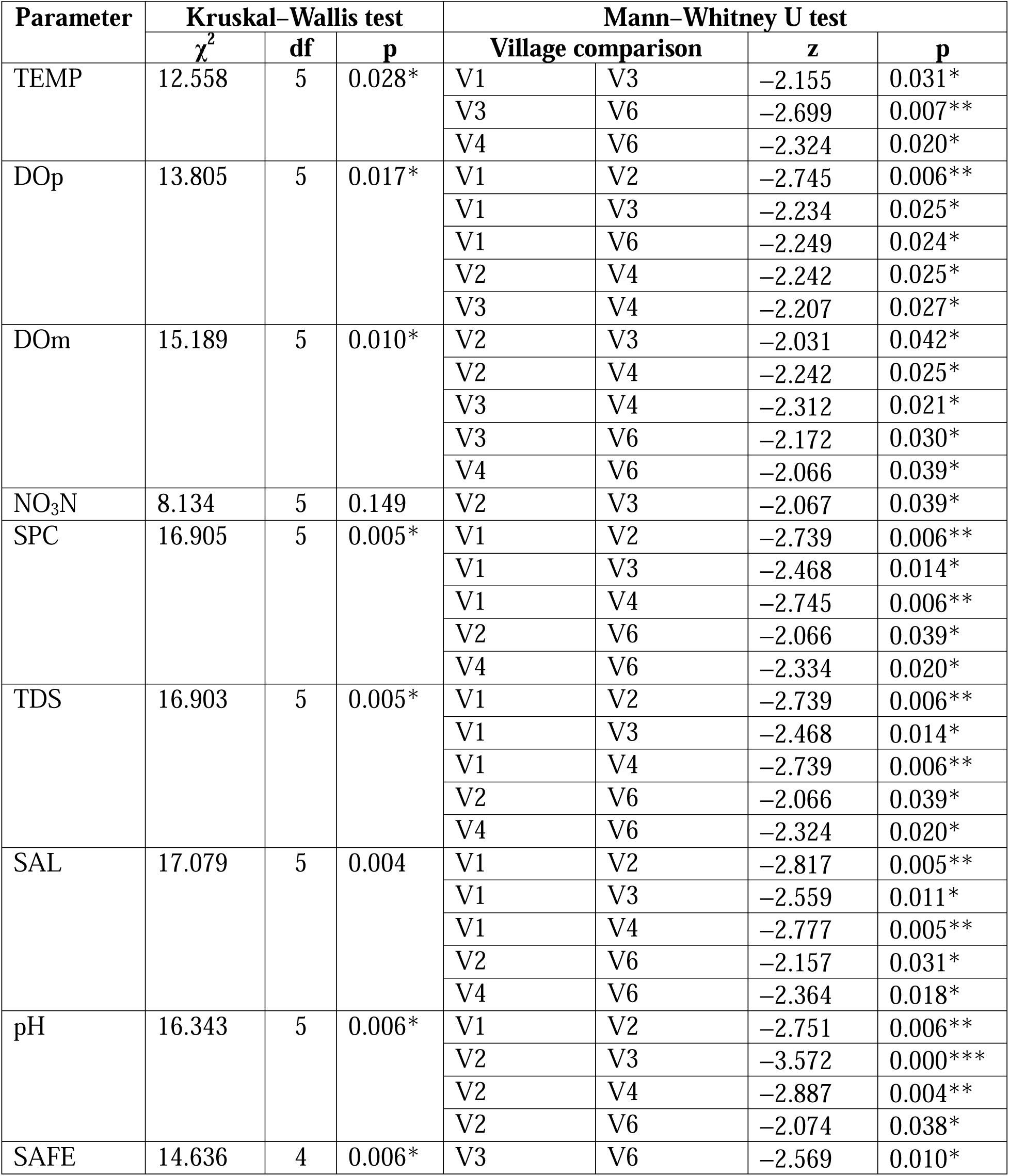
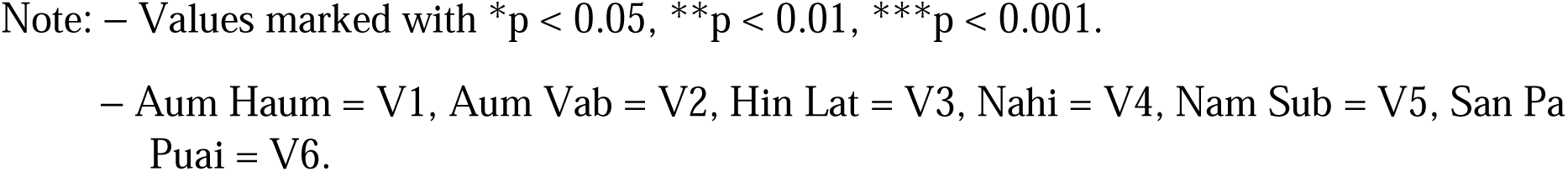
Comparison of water quality parameters across villages using Kruskal–Wallis test and post hoc Mann–Whitney U tests.

The PCA biplot illustrates the clustering of six villages, divided into two groups based on their water status, water quality parameters, and safety levels: a risk group (purple dotted circle) and a safe group (green dotted circle) (Fig. 4). The risk group, represented particularly by water sources from all villages, is characterized by higher loadings of NO3N, pH, TEMP, and TOX. This group is positioned opposite to the directions of FLOW and SAFE in the biplot, which indicates that it might involve unsafe, stagnant water sources. Unfortunately, the water sources in this group are often used for drinking, suggesting a potential health risk. The findings for the risk group indicate that residents using water sources in this group might be exposed to pesticides via their water and consume water of low quality.

**Fig. 4.**
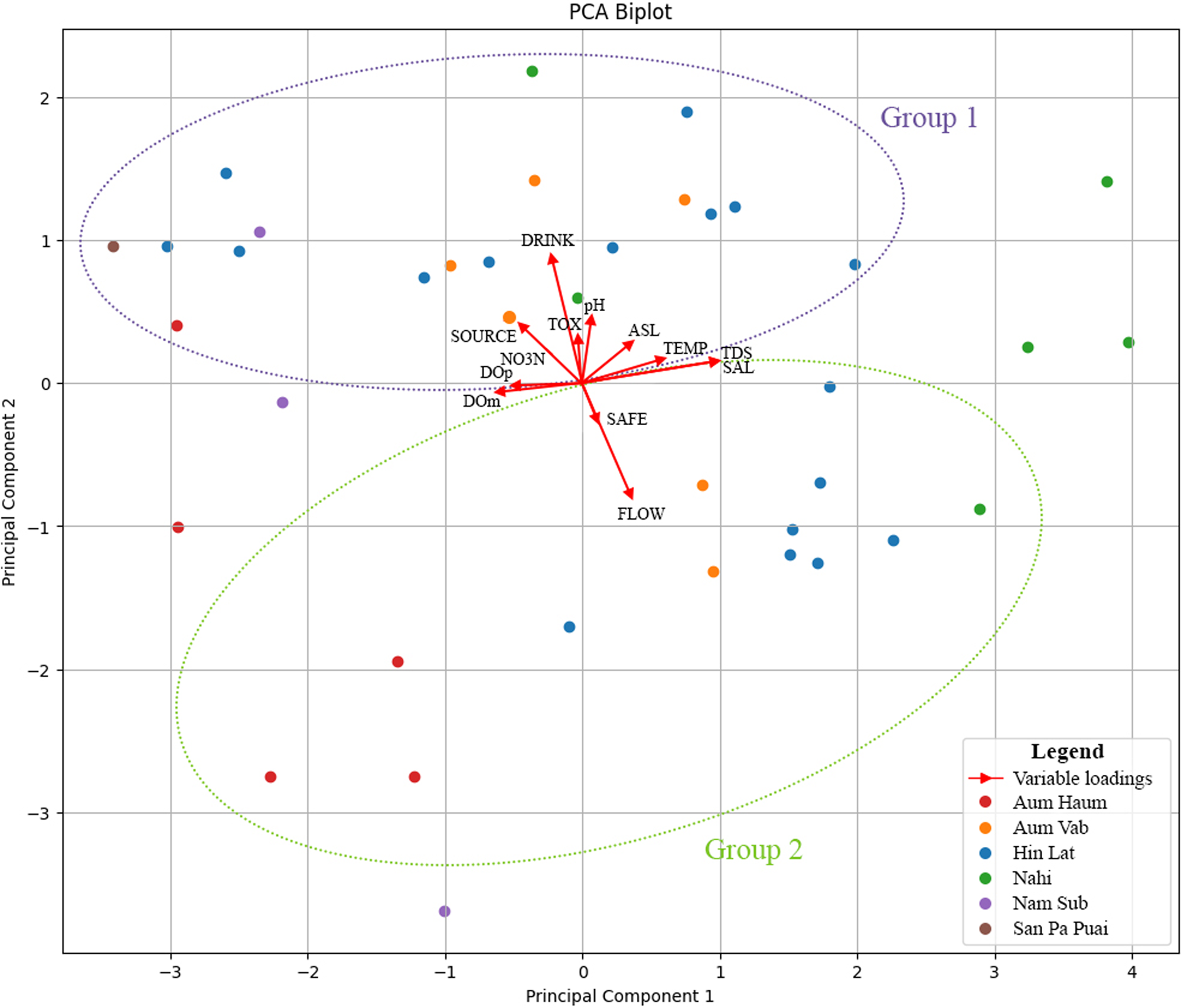
PCA biplot based on data from 38 water sources assessed across six villages upstream of Bhumibol Dam, between 19 and 21 November, 2024. Variables include source (SOURCE), drinking status (DRINK), flow condition (FLOW), height above sea level (ASL), specific conductivity (SPC), nitrate nitrogen (NO3N), mass of dissolved oxygen (DOm), percentage of dissolved oxygen (DOp), pH, salinity (SAL), temperature (TEMP), presence of pesticides (TOX), and toxicity levels (SAFE).

Conversely, the safe group, including sites such as Aum Haum, Aum Vab, Hin Lat, and Nahi, exhibited clustering associated with variables such as FLOW and SAFE, indicating that these sites tend to have safe, flowing water with low loading of pH, NO3N, and TOX. However, in the PCA biplot, this group is positioned away from the SOURCE and DRINK vector directions, suggesting that the residents of these villages may prefer not to drink water from these natural sources , and instead use it primarily for activities such as cooking and cleaning. Overall, this PCA plot effectively profiled water sources based on key water quality parameters, relating specific site conditions to the ecological and health risks that they pose.

### Pesticide contamination in the environment

In addition to assessing water sources, soil and agricultural products were collected and analyzed using the GT-Pesticide Residual Test Kit to determine contamination and toxicity levels. This kit specifically detects organophosphate and carbamate compounds. In this study, seven soil samples were collected across four villages: Aum Haum (3 samples), Aum Vab (1 sample), Hin Lat (1 sample), and Nahi (2 samples) (Table 3). The results indicated that all samples were contaminated with pesticides; however, the contamination levels remained within safe limits.

**Table 3.**
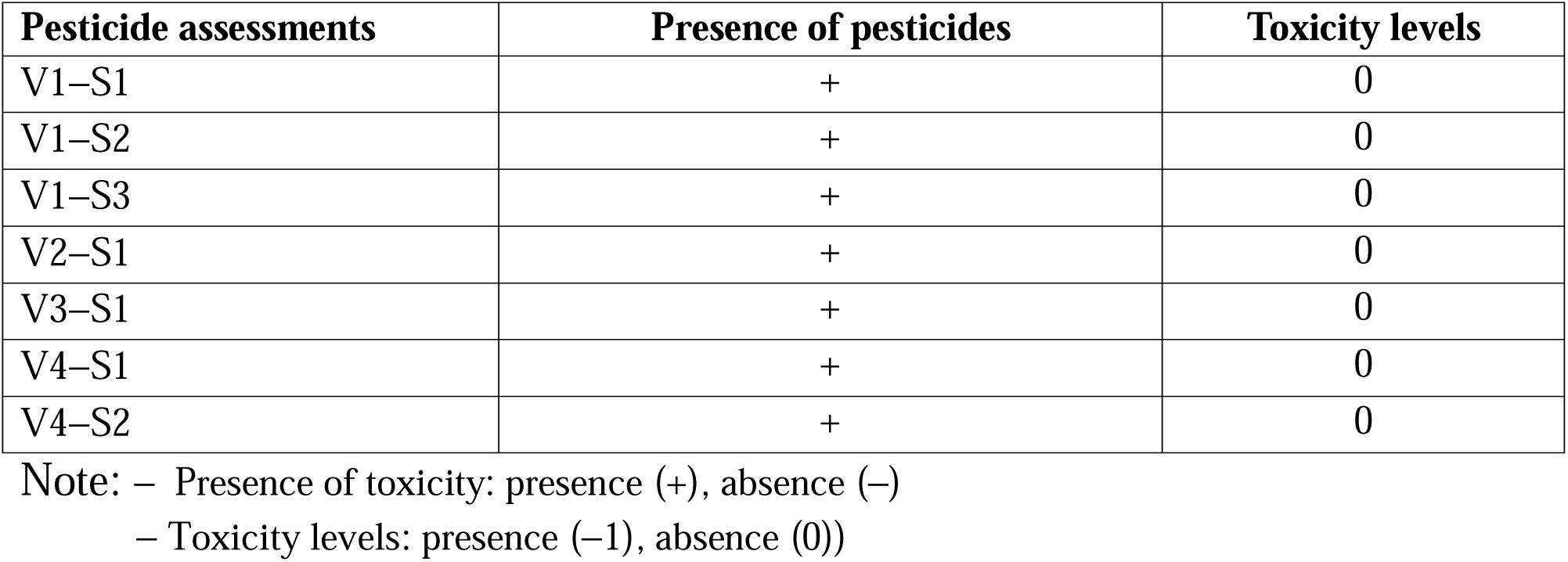
Assessment of presence and toxicity levels of pesticides in soil across four villages. S = sample.

Regarding agricultural products, various edible parts of crops were collected from the same sites as the soil samples across the four villages. Most edible plant parts showed no detectable pesticide contamination, especially those above ground (Table 4). However, three plants, arrowroot (*Canna edulis*), lemongrass (*Cymbopogon citratus*), and taro (*Colocasia esculenta*), from Hin Lat (V3) had edible below-ground parts that contained pesticide residues. Despite this, all samples remained within safe levels (Fig. 5).

**Fig. 5.**
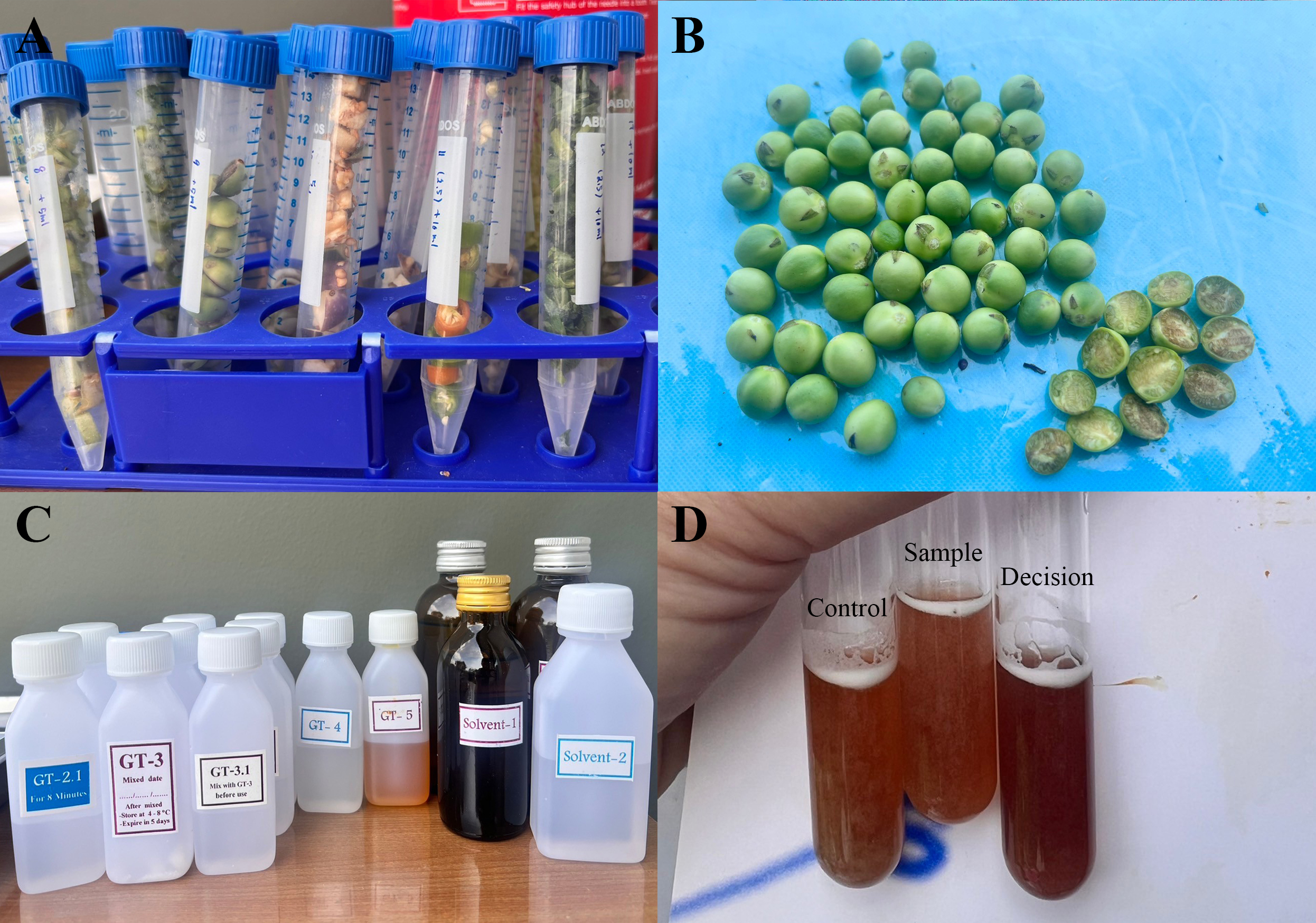
Analysis of agricultural products for pesticide contamination: (A, B) agricultural products; (B) turkey berry (*Solanum torvum*); (C) GT-Pesticide Residual Test Kit; and (D) results demonstrating that the sample tube is safe, indicated by its colour being similar to that of the control tube and lighter than that of the decision (positive) tube.

**Table 4.**
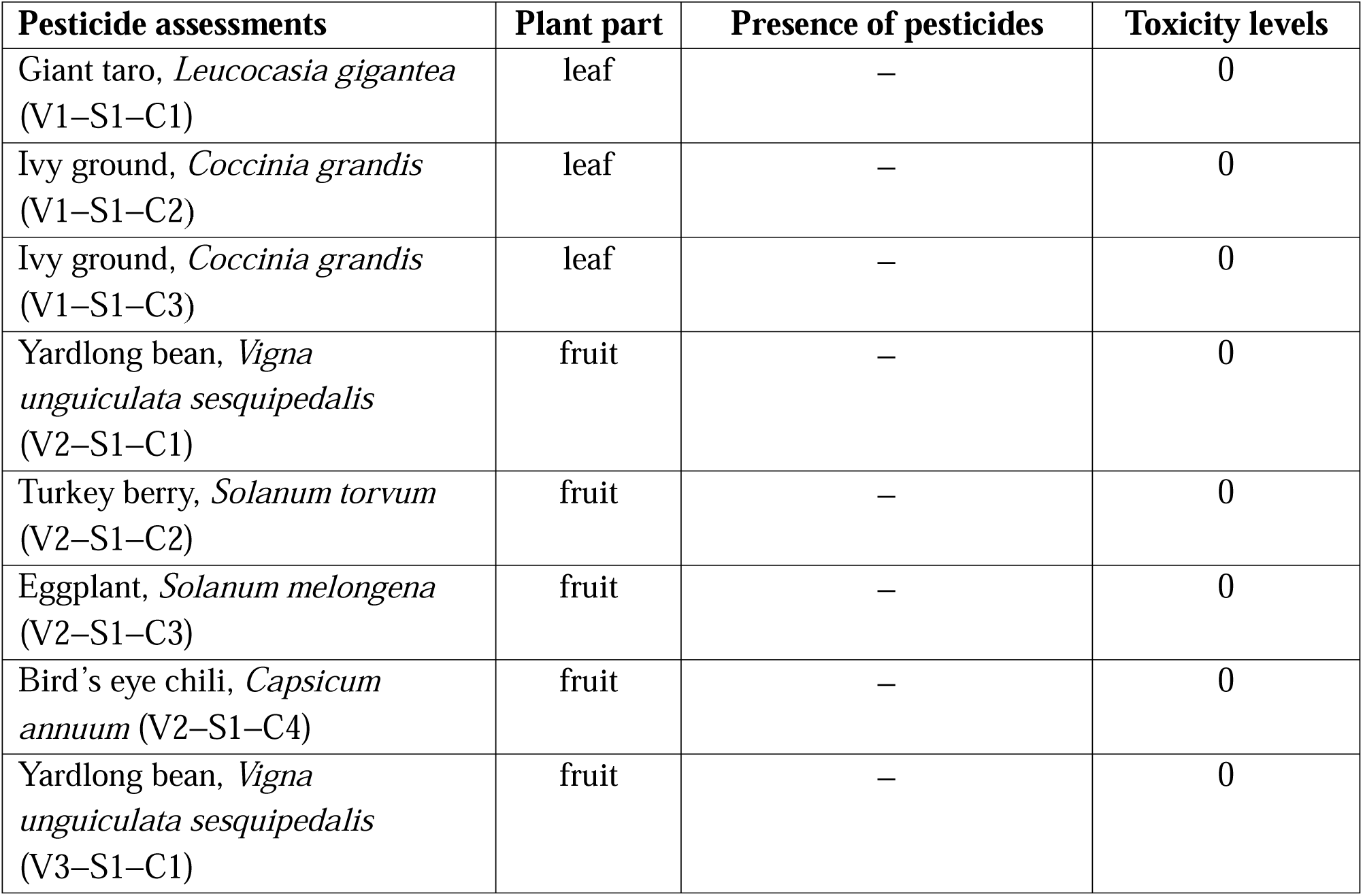
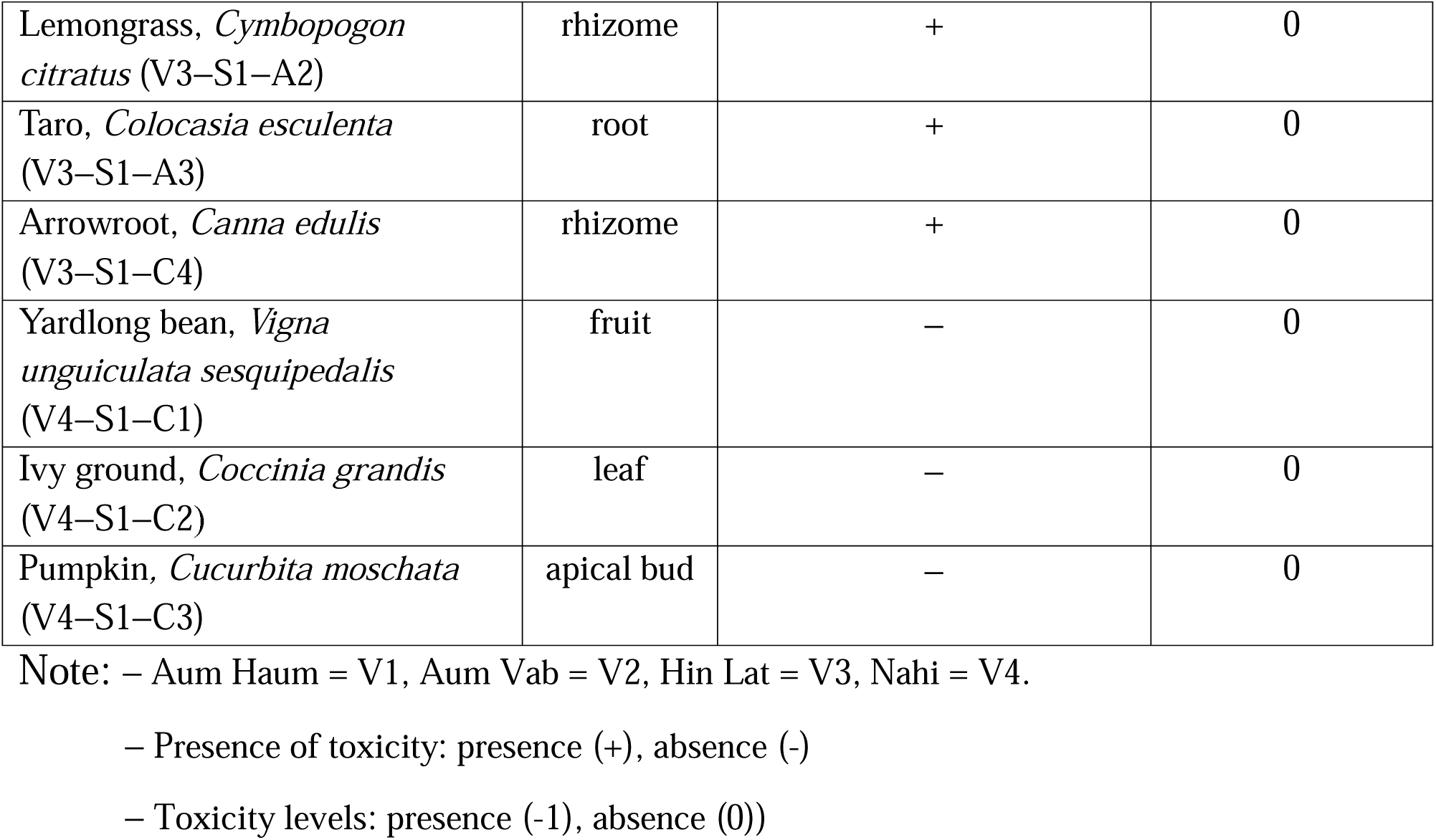
Assessment of presence and toxicity levels of pesticides in agricultural products, based on analyses of edible parts across four villages. S = sample, C = crop.

These results suggest that the soil in these villages is contaminated with pesticides, which may influence the plants growing there. This poses a potential health risk to residents who consume these crops, particularly the below-ground parts of them.

### Water filter efficacy

A water filter, installed at Hin Lat in 2018, is frequently used by residents of that village and its surroundings to obtain water for drinking and cooking. Water samples collected at that site before and after filtration were stored at 1–4°C and immediately transported to the Public Health Laboratory Division, Department of Health, Ministry of Public Health, Thailand, within 24 h for analysis. The analyses included assessments of apparent colour (Pt-Co), turbidity (NTU), pH at 25°C, TDS (mg/L), hardness as reflected by CaCO_3_ (mg/L), sulfate (mg/L), chloride (mg/L), nitrate as NO^−^ (mg/L), fluoride (mg/L), nitrite as NO^−^ (mg/L), iron (mg/L), manganese (mg/L), copper (mg/L), zinc (mg/L), lead (mg/L), total chromium (mg/L), cadmium (mg/L), arsenic (mg/L), and mercury (mg/L), as well as bacteria in the form of coliforms (MPN/100 mL) and *E. coli* (MPN/100 mL). Water samples were tested three times over a 6-month period at approximately 3-month intervals. Descriptive statistics based on the results of these analyses are presented in Supplementary Table 1. However, owing to the data not being normally distributed, a Wilcoxon signed-rank test was performed. The results indicated no significant differences in any of the analyzed water-related variables between before and after filtration, suggesting limited effectiveness of this water filter in reducing physical, chemical, microbial, and heavy metal contaminants (Fig. 6).

**Fig. 6.**
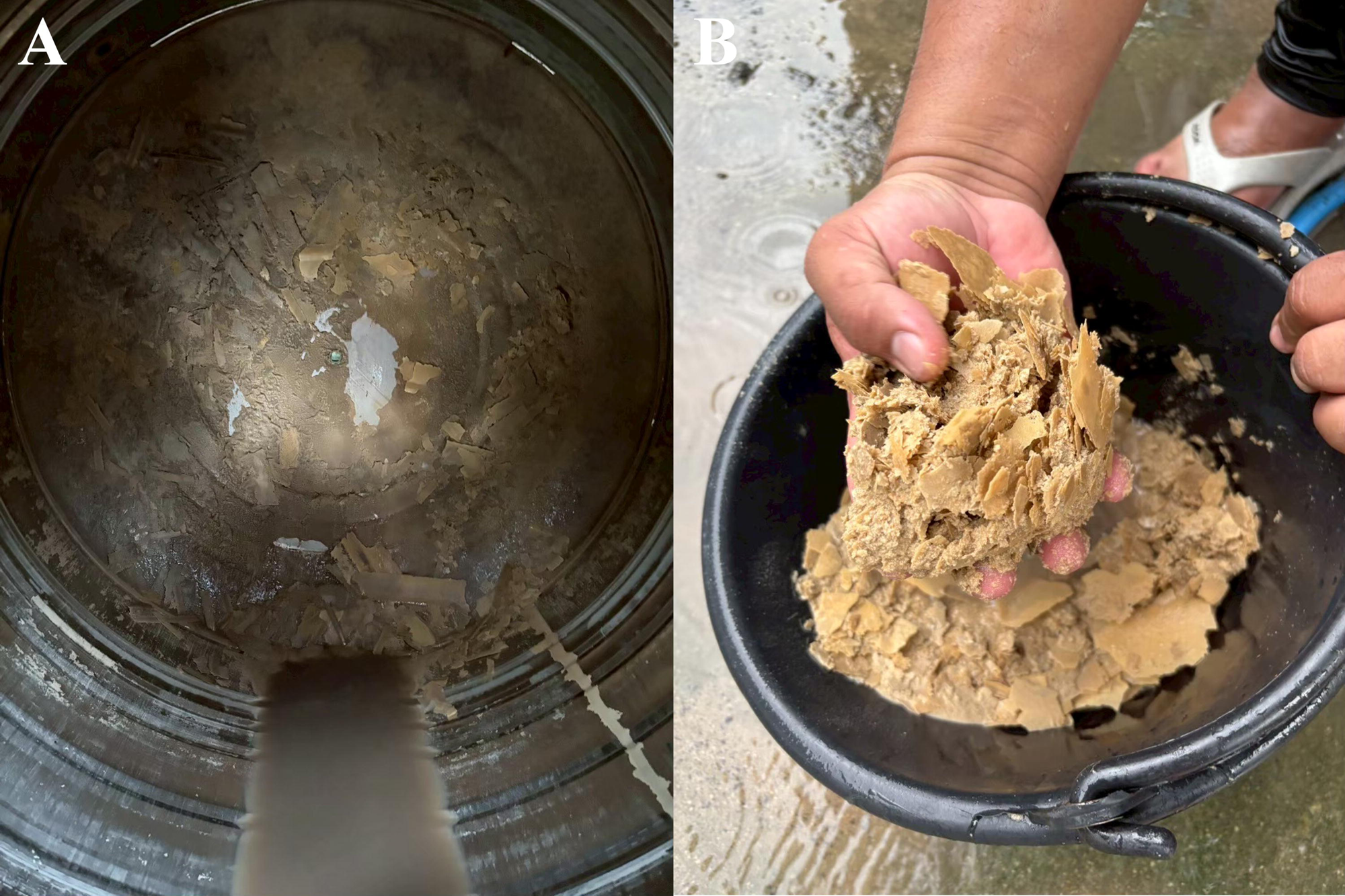
Accumulated sediment within a water filter tank.

## Discussion

### Patterns of water use in Thai mountain communities

Mountain ecosystems in Thailand, particularly the ranges like the Tenasserim Hills, are indispensable as regional headwaters, supplying perennial flow to various rivers and canals that sustain downstream lowland communities, evergreen nature of the mountainous regions year-round, and the upstream communities [22–26]. Mae Tuen Wildlife Sanctuary serves a similar critical function, acting as an upstream source for Mae Tuen Stream, which eventually feeds Ping River, a primary tributary of Bhumibol Dam. The people living in the catchment area upstream of the dam are entirely dependent on these localized, natural water sources within the sanctuary for their daily needs, including drinking, cooking, and cleaning.

The people of this region universally refuse to use or consume water directly from Ping River or the reservoir behind Bhumibol Dam. This is primarily because of the water’s high turbidity, which has led to the widespread perception that it is unclean. This aversion to highly turbid water is common among rural populations in Thailand and elsewhere who rely on surface water sources, where high sediment load is perceived as being strongly correlated with poor quality and biological contaminants [2,12,14,15,27–30]. Analysis of the patterns of water use confirmed that 34% of sampled sources were flowing natural water (mountain streams and waterfalls), which residents tend to favor for high-volume, non-drinking household activities such as washing and cleaning. This preference is rooted in the practical and perceived benefits of continuously moving water: its lower sediment load, reduced propensity for microbial contamination compared with stagnant pools, and an appearance suggestive of higher purity [2,29,30]. Furthermore, the ease of access to these sources of flowing water has allowed residents in places such as Aum Haum, Aum Vab, Nahi, and Nam Sub to establish settlements nearby. This in turn facilitates the transfer and storage of large volumes of water without the need to navigate kilometers of steep, difficult terrain to reach Bhumibol dam. However, despite this general preference for flowing water for use in everyday tasks like cooking and cleaning, the findings also raise concerns about the water used for drinking in this region, with still, sand-filtered puddles commonly being used for drinking water.

### Choice of drinking water and water quality heterogeneity

While the communities in this study exhibit a general preference for natural, flowing water sources for domestic uses such as cooking and cleaning, the analysis of drinking water sources revealed that more than half of the sampled drinking water (54.2% of drinking sources) was still water from dug sand-filtered puddles/storage containers (Fig. 2). This is particularly alarming because still or stagnant waterbodies, especially shallow, open pools such as these sand-filtered puddles (typically 30–60 cm in diameter and 5–10 cm in depth), inherently have a high risk of microbial contamination, exhibit increased concentrations of dissolved impurities, and can promote the transmission of vector-borne diseases [2,31–34].

The persistence of this practice appears to stem from a convergence of accessibility and indigenous knowledge. Generally, the sand-filtered puddles are easily dug close to residences, enabling the convenient collection of drinking water on a daily basis. More importantly, residents operate under the belief that the sand and soil naturally filter out “dirt, microbes, and toxins,” mimicking the principles of a slow sand filter (SSF) [35,36]. However, the efficacy of these rudimentary, unmanaged pools differs vastly from that of engineered SSFs, which require deep sand beds and a stable biological layer (dirty layer, *schmutzdecke*) to effectively remove pathogens [35–38]. The practice of using the same shallow puddle for 2–8 weeks before creating a new one risks the accumulation and possible fermentation of organic matter, leading to a potential increase in microbial load over time, creating a localized, ongoing health hazard [33,35,39,40].

Furthermore, statistical analysis supports the existence of significant heterogeneity in water quality across the six villages, as revealed by the Kruskal–Wallis test showing significant variations in SPC, TDS, and pH (Table 2). Specifically, villages V2 (Aum Vab), V3 (Hin Lat), and V4 (Nahi) exhibited significantly higher SPC and TDS, which aligns with the PCA risk-based clustering and indicates mineral-rich water conditions. In mountain hydrogeology, elevated SPC and TDS are directly indicative of high mineral content, predominantly in the form of alkaline minerals such as calcium and magnesium, which are dissolved as water percolates through the bedrock [41–43]. Water with this geochemical characteristic, while occasionally valued for its taste, might be directly linked to the health complication of bladder stone formation (urolithiasis), which is common in this region. In fact, according to community health reports, between 2019 and 2023, 22 people from Hin Lat reported having difficulty urinating (5 of whom received treatment for urinary tract stones). In line with this, studies have consistently demonstrated that consuming hard water (high in Ca^2+^ and Mg^2+^) can increase the concentration of stone-forming minerals in urine, significantly raising the risk of calcium oxalate stones, particularly in individuals with a genetic predisposition for them or inadequate hydration [12,33,34,44,45]. The differences in pH of the water obtained from the analyzed water sources among the villages in this study, such as the high mean pH of 8.32±0.10 at V2, further suggest that local geological or hydrogeological variations are the primary drivers determining the mineral composition, thereby generating distinct, site-specific health risks across the communities.

### Evidence of pesticide contamination and associated risk

The PCA provides strong empirical evidence that the water used by the communities in the studied region constitutes a critical environmental health hazard. The analysis effectively clustered certain water sources into a “risk group” defined by high factor loadings for (NO3N), alkaline pH, and TOX (Fig. 4). Alarmingly, many of the sources categorized into this cluster are those actively used by residents for daily drinking water. The strong correlation observed in the PCA between the NO3N and TOX vectors is a convincing indication of a common, diffuse source of contamination, which is clearly consistent with agricultural run-off (e.g., of agricultural chemicals) from non-point sources. This contamination pathway is further substantiated by findings from analyses of soil and water in the environment. Specifically, pesticide residues (organophosphate and carbamate compounds) were detected in all tested soil samples and within some collected water sources (Table 3). Critically, this contamination extended to the next trophic level, as evidenced by detectable levels of contaminants in the below-ground edible parts of perennial crops (arrowroot, lemongrass, taro) from V3 (Hin Lat) (Table 4). Although rice was not tested in this study because the work was performed outside of its growing season, conversations with residents showed that agrochemicals are used in paddy fields in the region.

These findings support the hypothesis that prevalent monoculture farming practices in the region, particularly agrochemical use in paddy fields, have resulted in the environmental partitioning and bioaccumulation of chemical pollutants, exposing residents through both their primary drinking water sources and their staple foods [46–48].

While the qualitative field kit determined the pesticide levels to be “safe,” this classification only reflects the kit’s detection limits and does not imply toxicological safety. Quantitative laboratory methods (e.g., gas chromatography-mass spectrometry (GC-MS)) are required to determine actual exposure levels. Nonetheless, the persistent detection of organophosphate and carbamate residues in water, soil, and crops signals a clear risk of chronic exposure. These compounds are known cholinesterase inhibitors, for which even low-level, long-term exposure, which is common in agricultural communities, has been linked to neurological, developmental, and endocrinological abnormalities [49–52]. Given the findings indicating such persistent chemical loading, there is an urgent need for quantitative assessments using laboratory-grade analytical methods in order to determine actual concentrations and better estimate the burden in terms of disability-adjusted life years (DALY).

Furthermore, the high NO_3_N levels observed in the risk group pose a separate and immediate toxicological threat. Excessive nitrate exposure can oxidize hemoglobin in the blood, leading to the severe condition of methemoglobinemia (or “blue baby syndrome”), which is particularly dangerous for infants and vulnerable populations [53–56]. The co-occurrence of these two major agricultural pollutants (NO3N and pesticides) in the communities’ preferred sources of drinking water underscores the existence of a compounded environmental health crisis exacerbated by inadvisable choices when selecting sources for drinking water and its inadequate treatment.

### Inadequate water infrastructure and public health implications

The community water infrastructure, specifically the filter system installed at Hin Lat, represents a profound shortcoming in terms of efforts to protect public health. In fact, specialized laboratory analysis of samples taken over a 6-month period demonstrated its complete lack of efficacy: There were no significant differences in physical, chemical, microbial, or heavy metal contaminant levels between the pre-filtration and post-filtration tanks. This directly supports the conclusion that the filter does not function effectively at removing contaminants and ensuring a supply of clean, potable water. This failure is doubly concerning, because it was previously shown that the water supply supposedly improved by the installation of this filter actually contained high levels of coliform bacteria (260 MPN/100 mL), including fecal ones (7 MPN/100 mL) far exceeding national and WHO safety levels for drinking water [2,55]. The sustained presence of pathogens coupled with the ineffective filtration system means that the community is not only relying on contaminated water but also under the misapprehension that its water supply is safe.

The confluence of this failed infrastructure and risky water-related practices create a compounded health burden for the Paganyaw communities. Specifically, people living in this region are simultaneously subjected to a double exposure risk. On the one hand, they risk acute exposure to microbial pathogens from the ongoing presence of fecal coliform bacteria in their water sources, leading to conditions such as diarrhea and gastrointestinal illnesses [33,35,40,41]. On the other hand, they suffer chronic exposure to alkaline minerals (linked to urolithiasis) and pesticide nitrate residues (linked to long-term systemic and neurological issues) because of their reliance on still sand-filtered puddles as sources of drinking water [12,33,34,44,45,53–56].

This situation highlights the urgent need for effective, sustainable water management in isolated, mountainous parts of Thailand like that upstream of Bhumibol Dam. Merely installing infrastructure to provide clean water, without subsequent technical validation, rigorous maintenance, and robust water safety plans, is insufficient to ensure water security in remote, vulnerable mountain communities. The findings of this work reveal the need for a shift toward context-appropriate, decentralized water treatment solutions that are locally manageable and resilient to the environmental and operational challenges of particular regions [57].

## Declaration of generative AI and AI-assisted technologies in the writing process

During the preparation of this work, the authors used ChatGPT and Gemini in order to check the grammar and improve the language of the manuscript. After using these tools, the authors reviewed and edited the content as needed and took full responsibility for the content of this manuscript.

## Data Availability

All data produced in the present work are contained in the manuscript

## Acknowledgment

This research project received funding support from Chulabhorn Royal Academy. Access to YSI EXO multiparameter instruments was supported by the Department of Zoology, Faculty of Science, Kasetsart University. Additionally, we thank Tom Buckle from Scribendi (www.scribendi.com) for editing a draft of this manuscript.

## Supporting information

**Supplementary Table 1.**
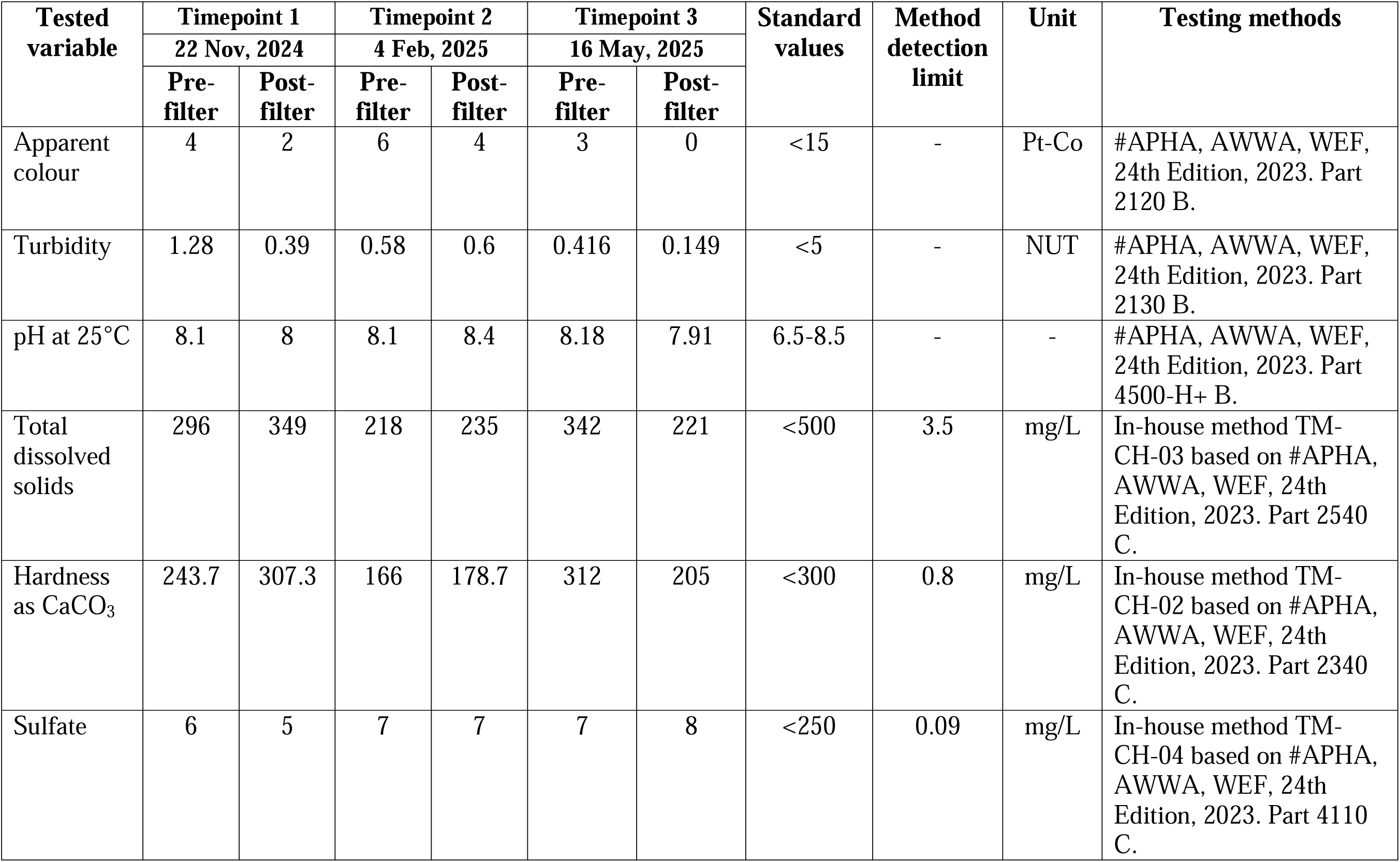
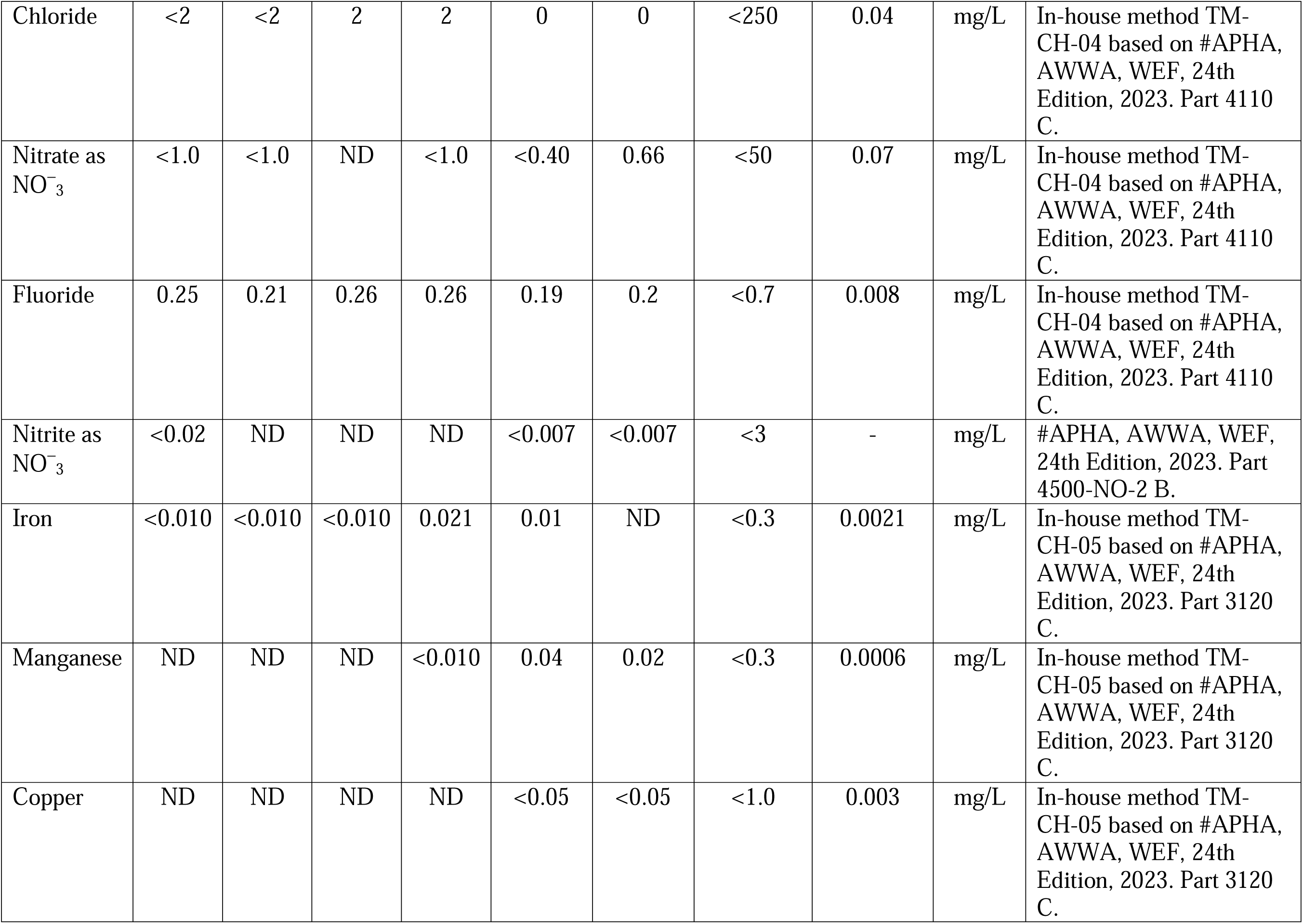
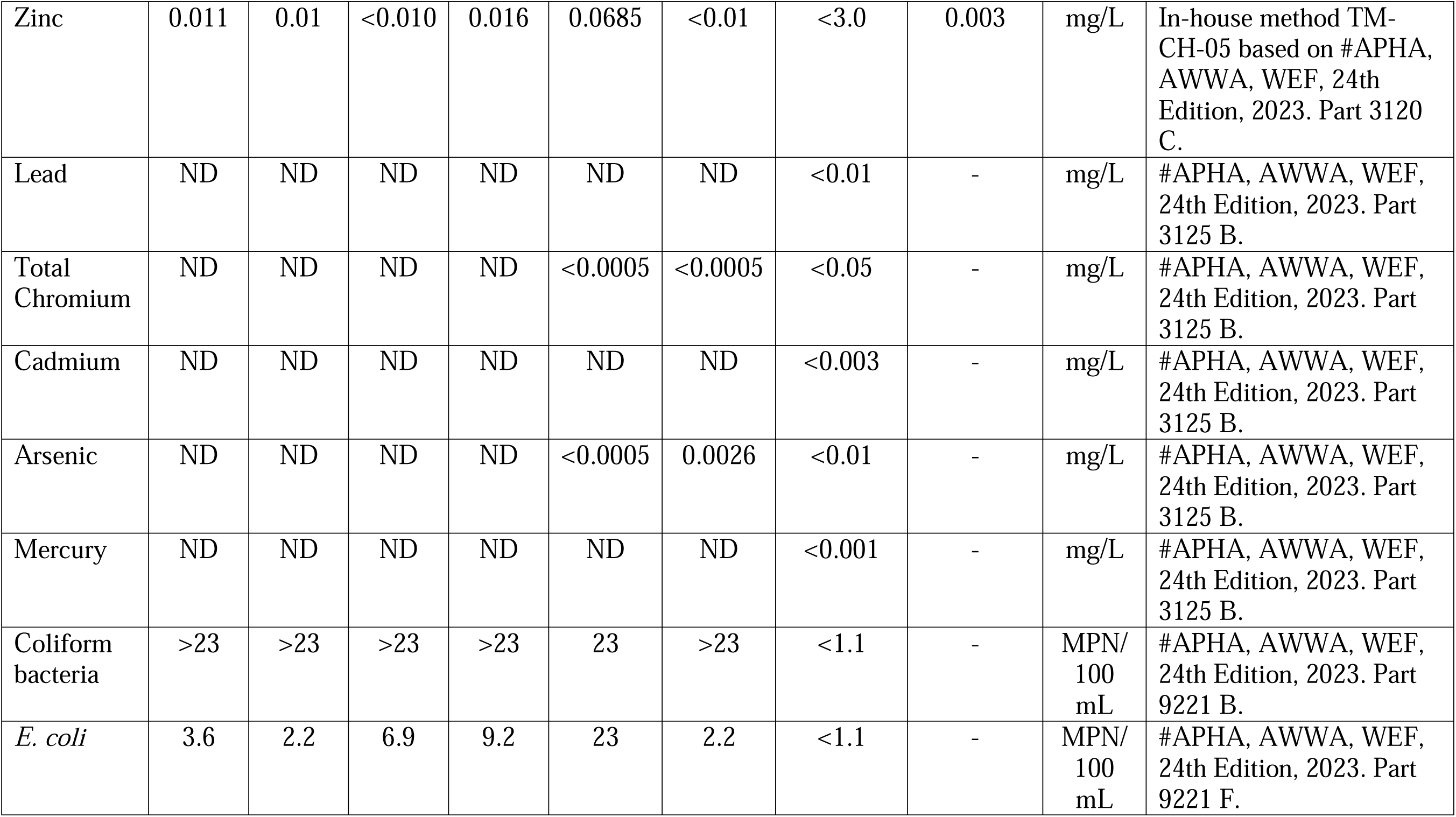
Descriptive analysis of water quality parameters of samples from a community water filters (Village V3, Hin Lat), evaluated across three sampling points over 6 months at approximately 3-month intervals.

